# Local socioeconomic status and paramedic students’ academic performance

**DOI:** 10.1101/19008516

**Authors:** Lydia Hamel, Ashley Procum, Justin Hunter, Donna Gridley, Kathleen O’Connor, Thomas Fentress, Christopher Goenner, Sahaj Khalsa, Alan M. Batt

**Affiliations:** Regions Hospital, St. Paul, Minnesota; Public Safety Group, Burlington, Massachusetts; Oklahoma State University; PhD candidate, Department of Paramedicine, Monash University, Victoria, Australia; UCLA Center for Prehospital Care, Los Angeles, California; Methodist Hospitals, Gary, Indiana; EMS, Central Piedmont Community College, North Carolina; Emergency Medical Services Institute, Santa Fe Community College, New Mexico; Fanshawe College, Ontario, Canada; Paramedic Science Discipline, CQUniversity, Queensland, Australia; Department of Paramedicine, Monash University, Victoria, Australia

## Abstract

Research indicates students of lower socioeconomic status (SES) are educationally disadvantaged. We sought to examine differences in paramedic student academic performance from counties with varying SES in the United States. Student performance data and SES data were combined for counties within the states of California, Mississippi, Louisiana, Texas and Virginia. Linear multiple regression modelling was performed to determine the relationship between income, high school graduation rate, poverty and food insecurity with first-attempt scores on the Fisdap Paramedic Readiness Exam (PRE) versions 3 and 4. Linear regression models indicated that there was a significant relationship between county-level income, poverty, graduation rate, food insecurity, and paramedic student academic performance. It remains unclear what type of relationship exists between individual SES and individual academic performance of paramedic students. These findings support the future collection of individual student level SES data in order to identify issues and mitigate impact on academic performance.

## Introduction

With over 50 million Americans living at or below the poverty line (Oxfam, 2019), poverty is a social issue that is endemic across the United States. Those living in poverty face challenges in accessing healthcare, education and food – things many take for granted. In addition, over 50 million Americans are food insecure, meaning they lack consistent access to enough food to lead an active, healthy life (Gundersen, 2013). This results in children and students going to bed hungry at night. Often times, a school breakfast will be their only meal for the day. Living in poverty means these students are less likely to finish high school or college, which may further influence their lifestyle as adults, including job prospects, decisions around diet, recreational activities, and substance use (Lacour & Tissington, 2011; Ladd, 2012). In addition, poverty and low socioeconomic status (SES) are associated with visible low social status, and this can be a source of intrinsic chronic stress, increasing vulnerability to distress and potentially destructive coping mechanisms (Mulia, Schmidt, Bond, Jacobs, & Korcha, 2008).

SES, which is a measure based largely on income, education and occupation, is a major social basis for inequalities and an important predictor of an individual’s health, career and lifetime earnings. Low SES is strongly associated with lower educational attainment (Lacour & Tissington, 2011; van der Berg, 2008), and this association is observable across data from local to national levels (Ladd, 2012). Students from low SES households are more likely to experience food insecurity (Dean & Sharkey, 2011), which is strongly associated with precarious housing status (Goldrick-Rab, Richardson, Schneider, Hernandez, & Cady, 2018; Hughes, Serebryanikova, Donaldson, & Leveritt, 2011; Silva et al., 2017), and in turn is related to lower academic success (Roustit, Grillo, Martin, & Chauvin, 2015). An estimated 25% to 36% of post-secondary students suffer from food insecurity (Silva et al., 2017), more so in the community college population (Goldrick-Rab et al., 2018). If students are food insecure, they may face difficulties in their educational pursuits.

Considering that socioeconomic issues exert such influence on student performance, educators should remain aware of such issues within their student population. Educators who are unaware of these influences may be less likely to connect students to appropriate supports in order to foster success. Unless educators appropriately support students from low SES backgrounds, paramedic training programs risk precipitating a cycle that makes them less likely to enrol in college in the first instance, more likely to fail out, and less likely to have formal qualifications when compared to their peers from higher SES backgrounds (Blanden & Gregg, 2004; National Student Clearinghouse, 2016). Lack of support ultimately negatively affects adult working life, in terms of both income and working hours (Duncan, Magnuson, Kalil, & Ziol-Guest, 2012; Duncan, Magnuson, & Votruba-Drzal, 2017)

Importantly, this means support both from inside and outside of educational institutions is essential. If students lack adequate support at home from family and peers, this can present a further barrier to their educational attainment (Engle, 2007). Due in part to the parents’ own education level, and the chronic stress associated with poverty and low SES, families living in high poverty, high unemployment and low education neighbourhoods are likely to employ fewer education-oriented practices with their children (Greenman, Bodovski, & Reed, 2011). Evidence shows that increased parental involvement in academic matters can mitigate some of the effects of low SES (Greenman et al., 2011). Increased family support has been demonstrated to directly influence overall academic achievement and retention rates at the community college level (Kuh, Kinzie, Buckley, Bridges, & Hayek, 2006).

Many paramedic programs are delivered through the community college system and therefore it is a source of concern that many paramedic programs may not routinely collect or use the availble information related to student SES. While the relationship between individual SES and academic performance in paramedic students remains unclear, there is strong evidence from the broader literature to suggest that low SES can result in negative effects on academic performance. Paramedic education programs need to collect and use this data, otherwise educators will remain blind to the effects of SES on students. Additionally, students from certain geographical areas may face challenges when they enter and paramedic education programs due to the local socioeconomic climate. It is highly unlikely that all students from a low SES background would report such issues, due to the significant stigma associated with poverty and low SES.

In light of the ease of availability of county-level SES data, the authors proposed to investigate if these data could be used to uncover some of the concerns highlighted in relation to poverty, income, food insecurity and parental education, specifically in relation to paramedic students. These insights will then be used to explore potential solutions for consideration.

### Aim

This study sought to examine the relationship between paramedic student academic performance and county-level SES indicators. Additionally, the authors sought to examine the relationship between a paramedic applicant’s parental education level and the applicant’s entrance exam scores. It was hypothesized that paramedic students from counties with lower SES (as defined by median income, percent living below the poverty line, percent of food insecurity and high school graduation rate) would demonstrate lower performance on exams when compared to students from counties with higher SES (as defined by the same criteria). It was also hypothesized that lower parental education level would be associated with lower applicant entrance exam scores.

## Methods

### Ethics approval

This study was conducted with ethics approval from Inver Hills Community College in Grove Heights, Minnesota.

### Data collection

Data were acquired for paramedic students with accounts in Fisdap™, an internet-based EMS education administrative database in the states of California, Virginia, Louisiana, Texas and Mississippi. These states were selected to allow for variation in SES as well as an adequate number of paramedic programs that enter data into Fisdap™. In addition to basic demographic data, entrance exam (EE) scores from September 2018 to January 2019, were collected for these students. Parent education level was also collected from EE data. Paramedic Readiness Exam (PRE versions 3 and 4) scores from 2011 to 2019, were obtained for these students as well. Only scores from the first attempt at the PREs were counted. Publicly accessible SES data for all counties within the identified states were acquired from the Robert Wood Johnson Foundation County Health Rankings for 2017, and US Census data. ZIP code data were converted to county for each state via an online US ZIP Code lookup tool.

### Inclusion and exclusion criteria

Records from students who had indicated their consent for research, had complete demographic information, including ZIP code, and who had made at least one attempt on the EE, PRE3 or PRE4 (not all programs use all exams) were included in the analyses. Records from students who did not give consent for research, that had incomplete demographic data, or missing ZIP code information were excluded from analyses. Only first attempt scores were included, and counties with less than 5 reported scores were excluded.

### Data analysis

Statistical analyses were performed in R (R Core Team, 2019). A multiple linear regression analyses was performed utilizing PRE3 and PRE4 scores, combined with income, poverty level, high-school graduation rate and food insecurity data to determine predictive relationships between these variables. To obtain the relationship of each parental educational level EE score, a one-way analysis of variance (ANOVA) was conducted with score as the dependent level and the parent education level (with six levels) as the factor. Continuous variables are presented as means, median, standard deviations (SD). p <0.05 was considered statistically significant.

### Definitions

The following definitions are utilized in the County Health Rankings Report (Robert Wood Johnson Foundation, 2019), and the US Census Data (United States Census Bureau, 2019), and are applicable to the data and discussion in this study.

- Socioeconomic status (SES): an economic and sociological combined total measure of a person’s work experience and of an individual’s or family’s economic and social position in relation to others, based on income, education, and occupation.
- Poverty: the Census Bureau uses a set of money income thresholds that vary by family size and composition to determine who is in poverty. If a family’s total income is less than the family’s threshold, then that family and every individual in it is considered in poverty. The official poverty thresholds do not vary geographically, but they are updated for inflation. The official poverty definition uses money income before taxes and does not include capital gains or noncash benefits (such as public housing, Medicaid, and food stamps). [variable: Poverty]
- Food insecurity: the percentage of the population who did not have access to a reliable source of food during the past year. This measure was modelled using information from the Community Population Survey, Bureau of Labor Statistics, and American Community Survey [variable: Food]
- High school graduation: the percentage of ninth grade cohort that graduates high school in four years. [variable: GradRate]
- Median household income: the income where half of households in a county earn more and half of households earn less. Income, defined as “Total income” is the sum of the amounts reported separately for: wage or salary income; net self-employment income; interest, dividends, or net rental or royalty income or income from estates and trusts; Social Security or Railroad Retirement income; Supplemental Security Income (SSI); public assistance or welfare payments; retirement, survivor, or disability pensions; and all other income. [variable: Income]

## Results

### PRE3 Data

There were 3,697 records from the five states. These were aggregated to calculate the mean score and number of results by county. The final data set used for analyses (counties >5 reported scores) consisted of data from 151 counties.

### PRE4 Data

There were 1,293 records from the five states. These were aggregated to calculate the mean score and number of results by county. The final data set used for analyses (counties >5 reported scores) consisted of data from 60 counties.

### EE Data

There were 3,607 records from students who took the entrance exam between September 2018 and January 2019. Table 1 outlines the characteristics of included data (number of records, mean score, number of counties, etc).

**Table 1.** Characteristics of included data.

| Exam | n<br>(records) | Mean<br>score | SD | n<br>(counties) | Mean<br>reports<br>per<br>county | SD | Range |
| --- | --- | --- | --- | --- | --- | --- | --- |
| PRE3 | 3697 | 69.76 | 9.13 | 151 | 20.25 | 28.4 | 5-237 |
| PRE4 | 1293 | 72.1 | 7.2 | 60 | 14.68 | 12.38 | 5-77 |
| EE | 3607 | 79.26 | 8.65 | - | - | - | - |

A linear multiple regression model was conducted to determine the relationship of each SES variable to PRE3 score. The model was *PRE3 Score ∼ Income + GradRate + Poverty + Food*. The results of the regression are presented in Table 2. Results indicated there was a significant collective relationship between income, poverty, graduation rate, food insecurity, and PRE3 scores, (F(4,143)=10.66, p < .001, R2=.23). The individual predictors were examined further and indicated that income (t = 5.36, p < 0.001), graduation rate (t = −3.01, p < 0.01), poverty (t = 2.28, p < 0.05) were significant predictors in the model. Figure 1 demonstrates the relationship of each of the four SES variables with the PRE3 score. The y-axis measures the PRE3 score, while the x-axis measures the individual predictors.

**Table 2.** Results of linear multiple regression model for PRE3 data. *(***=p<0*.*001; **=p<0*.*01; *=p<0*.*05). Residual standard error: 3*.*771 on 143 degrees of freedom (3 observations deleted due to missingness) Multiple R-squared: 0*.*2296, Adjusted R-squared: 0*.*2081, F-statistic: 10*.*66 on 4 and 143 DF, p-value: <0*.*0001*.

| Variable | Est | Std. Error | t-value |
| --- | --- | --- | --- |
| Intercept | 75.8 | 4.53 | 16.73*** |
| Income | 0.00 | 0.00 | 5.36*** |
| High School<br>Graduation Rate | -0.14 | 0.05 | -3.01** |
| Poverty | 0.11 | 0.05 | 2.28* |
| Food insecurity | -0.16 | 0.1 | -1.62 |

**Figure 1.**
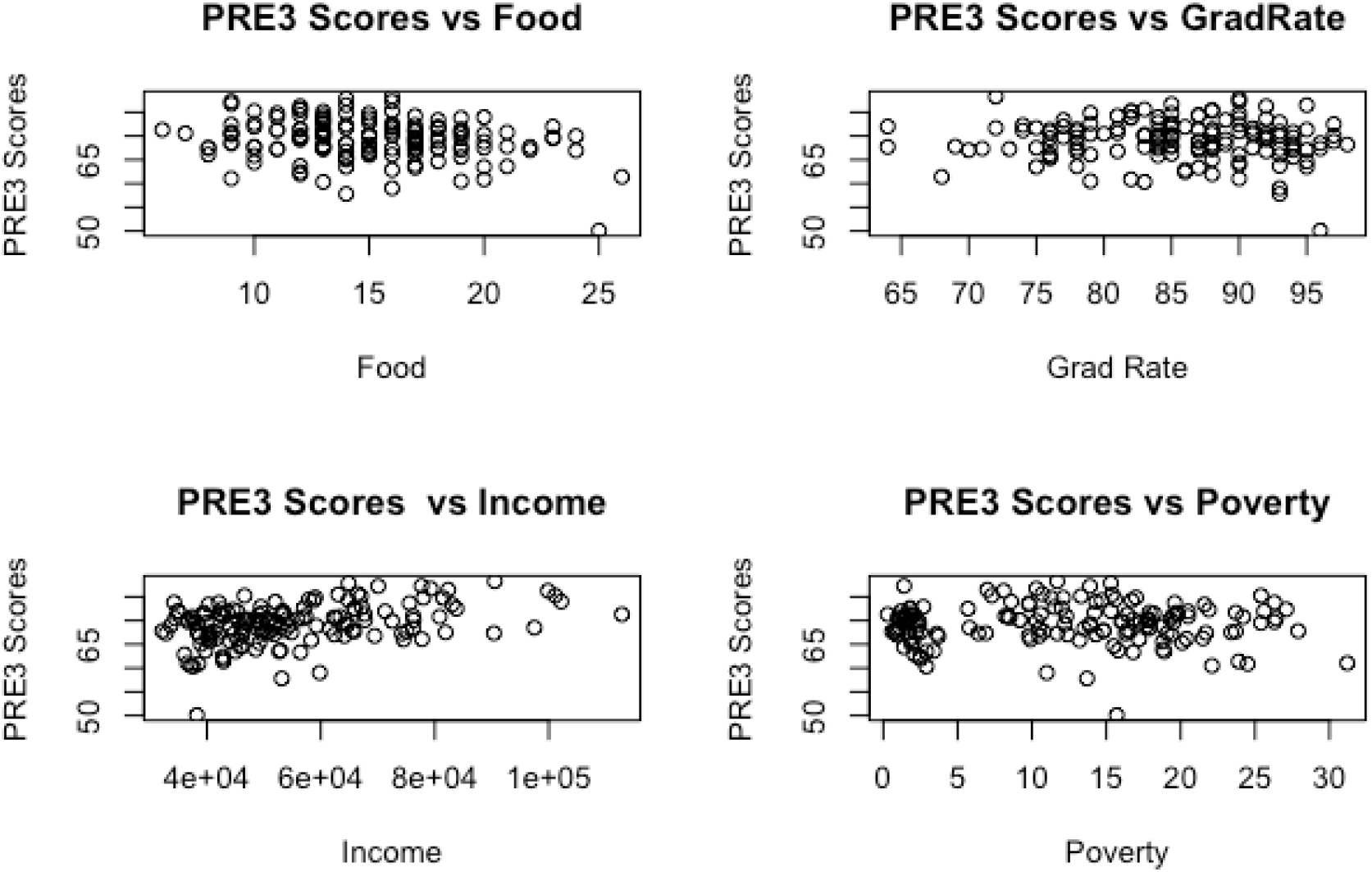
Scatterplots of SES and PRE3 data.

To determine the relationship of each SES variable to PRE4 score, a linear multiple regression model conducted. The model was *PRE4 Score ∼ Income + GradRate + Poverty + Food*. The results of the regression are presented in Table 3. These results indicate that there was a collective significant effect between income, poverty, graduation rate, food insecurity, and PRE4 scores, (F(4,54)=4.72, p < 0.01, R2=.26). The individual predictors were examined further and indicated that income (t = 3.71, p < 0.001) was the only significant predictor in the model. Figure 2 shows the relationship of each of the four SES variables with the PRE4 score. The y-axis measures the PRE4 score, while the x-axis measures the individual predictors.

**Table 3.**
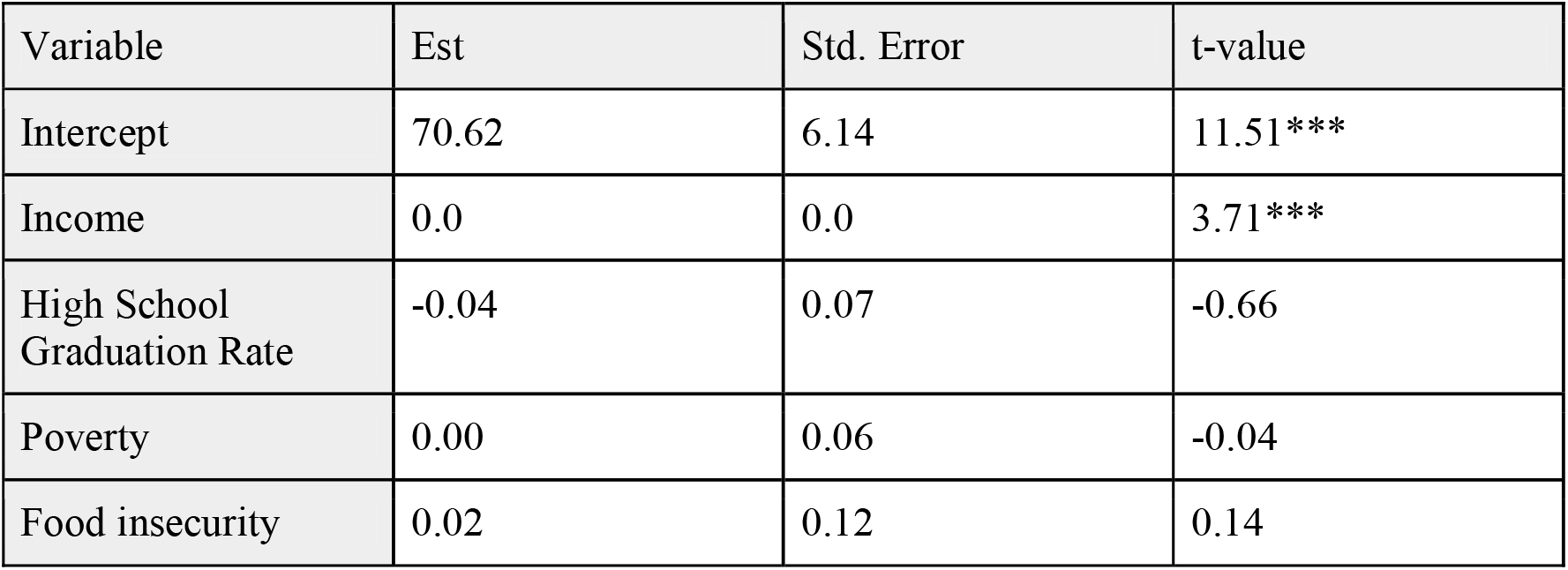
Results of linear multiple regression model for PRE4 data. *(***=p<0*.*001). Residual standard error: 2*.*841 on 54 degrees of freedom (1 observation deleted due to missingness), Multiple R-squared: 0*.*2592, Adjusted R-squared: 0*.*2043, F-statistic: 4*.*723 on 4 and 54 DF, p-value: <0*.*01*.

| Variable | Est | Std. Error | t-value |
| --- | --- | --- | --- |
| Intercept | 70.62 | 6.14 | 11.51*** |
| Income | 0.0 | 0.0 | 3.71*** |
| High School Graduation Rate | -0.04 | 0.07 | -0.66 |
| Poverty | 0.00 | 0.06 | -0.04 |
| Food insecurity | 0.02 | 0.12 | 0.14 |

Table 3. Results of linear multiple regression model for PRE4 data.
| Variable | Est | Std. Error | t-value |
| --- | --- | --- | --- |
| Intercept | 70.62 | 6.14 | 11.51*** |
| Income | 0.0 | 0.0 | 3.71*** |
| High School<br>Graduation Rate | -0.04 | 0.07 | -0.66 |
| Poverty | 0.00 | 0.06 | -0.04 |
| Food insecurity | 0.02 | 0.12 | 0.14 |

**Figure 2.**
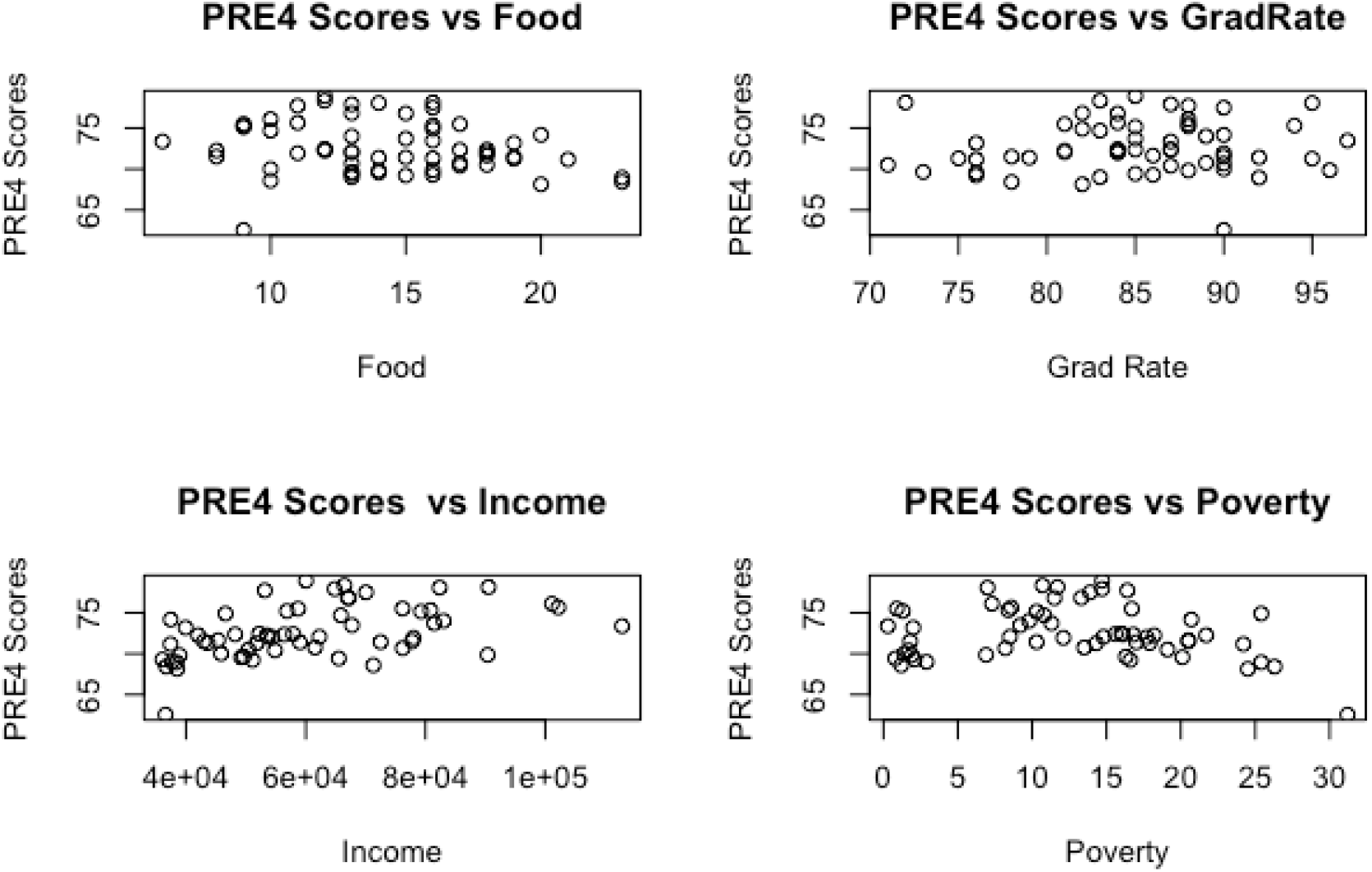
Scatterplots of SES and PRE4 data.

### Entrance exam

The boxplots in Figure 3 below show the distribution of entrance exam scores by parent educational level. The y-axis measures the EE score, while the x-axis measures the parent education level.

**Figure 3.**
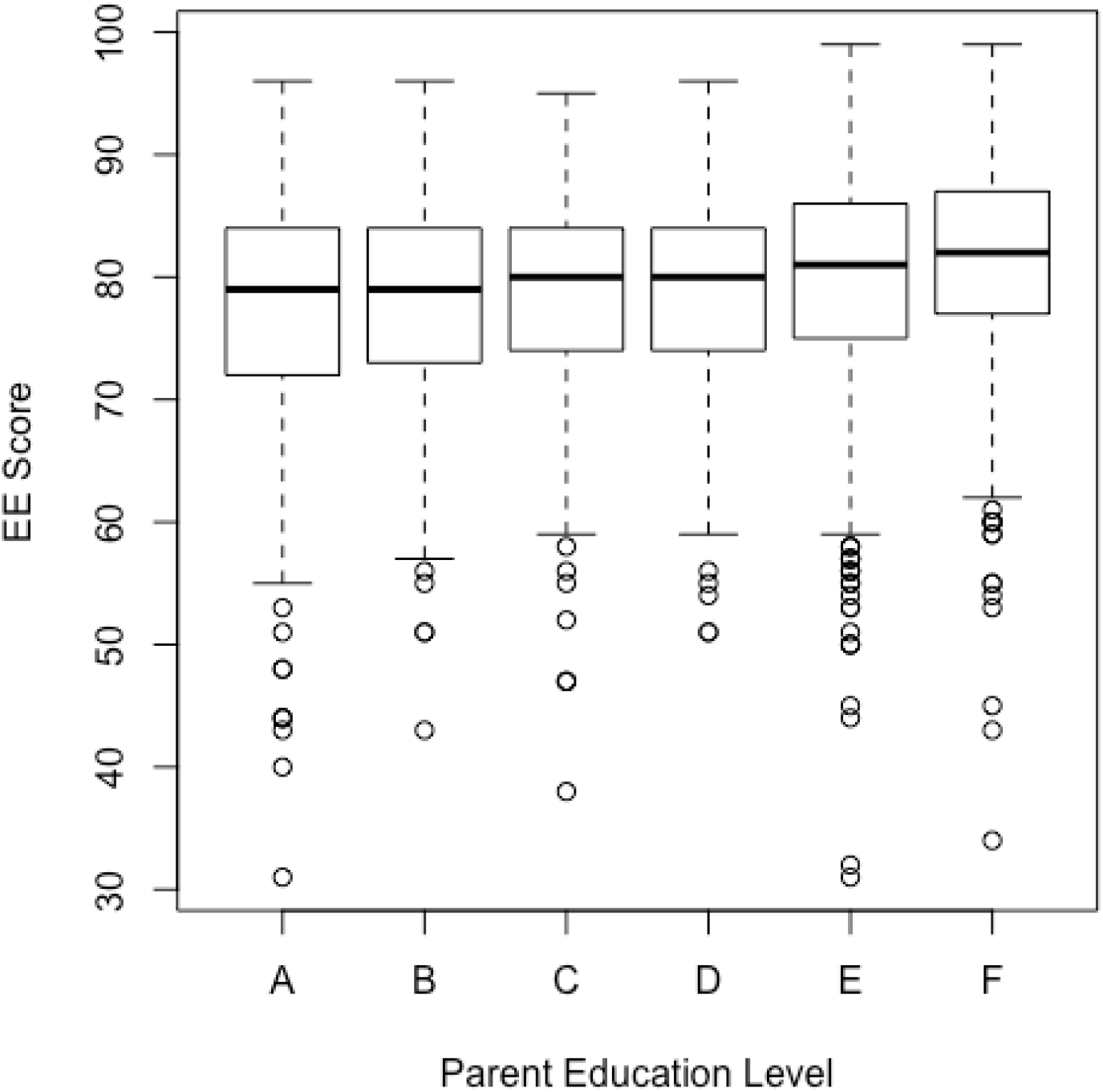
Boxplots of entrance exam scores by parent educational level. *A = High school diploma or less; B=Some college; C=Trade certification; D=Associate’s degree; E=Bachelor’s degree; F=Graduate degree (Masters, PhD)*.

The one-way ANOVA for parental level of education and EE score was statistically significant. (F(5,3601) = 18.23, p < 0.001). Students whose parents had a high school diploma or less had the lowest EE scores (Group A, mean = 77.42, SD = 9.21), while students whose parents had a graduate degree had the highest EE scores (Group F, mean = 81.55, SD =8.22).

## Discussion

This study demonstrated that county-level SES indicators, in particular a combination of income, poverty level, high school graduation rate and food insecurity, had a significant relationship with paramedic student academic performance. Median household income is a well-recognized indicator of income and poverty, and this income at a county-level was significantly related to exam scores in both PRE3 and PRE4 exams. With one in four children in the United States living on or below the poverty line (Oxfam, 2019), this is a cause for concern. Parental academic achievement was significantly associated with EE scores in this study. This is significant due to previous research which demonstrated that performance on the EE positively predicted first-time pass rate on the National Registry of Emergency Medical Technicians Cognitive Exam (Page, Bowen, & Stanke, 2015), and was associated with performance on cardiology and airway unit exams (Page, Bowen, & Stanke, 2013). EE performance was also associated with graduation success in paramedic programs (Renkiewicz et al., 2015).

There is significant stigma attached to lower SES (Johnson, Richeson, & Finkel, 2011; Stewart et al., 2009; Williams, 2009), which may present as a barrier to individual students seeking assistance. In order to remove such barriers, paramedic programs may consider interventions such as online resources regarding income support, the provision of scholarships, and access to confidential counselling and support services. In addition, programs should endeavour to highlight the availability of existing resources within the larger college or university community. These may include resources such as financial aid, health plans, legal advice, used book stores, bike-shares, and transit passes. Paramedic programs in the USA and Canada are often delivered through community colleges, and such students are more likely to go a day without eating than their university counterparts (Goldrick-Rab et al., 2018). Food insecurity and housing instability can negatively affect classroom attendance, academic performance and the ability to continue in higher education (Broton & Goldrick-Rab, 2016; Goldrick-Rab et al., 2018). Paramedic programs may need to consider implementing interventions such as food banks, sharing shops, breakfast clubs, information on food supplementation, and rent and meal subsidies where appropriate. Despite the evidence that SES affects student academic performance, paramedic programs in these jurisdictions do not routinely collect information on the individual SES of their students. It remains unclear what type of relationship exists between individual SES and individual academic performance of paramedic students. This study suggests that paramedic programs should attempt to collect individual SES data from students that can then be used to identify issues and mitigate any potential effects of SES on academic performance. Such data would need to be collected and acted upon in confidence.

### Limitations

The results presented are subject to several limitations. The study was limited to data obtained retrospectively, from self-reported demographic data, and first attempt at entrance, PRE-3 and PRE-4 paramedic exams submitted to Fisdap™. Not all paramedic programs in the United States use the Fisdap™ database and testing products. The sequence of tests, and the conditions under which the tests were conducted (proctored, open-book, timed etc.) cannot be determined from the Fisdap™ data. Students may have entered their program or work address instead of their home address. Counties and ZIP codes generally contain significant variability in SES, and this cannot be accounted for in our data. County-level data also do not reflect individual student SES, which is a more accurate predictor of individual student success. There is no prior evidence this data exists, hence the use of surrogate data markers. Counties that had less than 5 reported scores and records that were missing ZIP codes were excluded from analysis.

## Conclusion

This study has demonstrated that county-level SES factors are associated with paramedic student academic performance. While this study utilised county-level data as a surrogate for individual student SES, it does provide some insight into the potential effects that a student’s background may have on their academic performance. Accepting that the results need to be interpreted in the context of the significant limitations of such an approach, given the lack of other avenues by which to explore this issue, the authors believe that the results nevertheless improve the paramedic community’s understanding of the issue. The results of this study also serve to highlight for the first time within paramedic education the identified role of SES on academic performance. This issue remains (anecdotally) poorly addressed within paramedic education programs. Given the evidence that SES can impact academic performance in general, this study supports the collection of individual student level SES data in order to accurately identify and address its effects on student success in education programs.

## Data Availability

Available on request

## Acknowledgments

The authors would like to thank Public Safety Group, Jones and Bartlett Learning/Ascend Learning and Fisdap. Thanks to Dr. Adisack Nhouyvanisvong PhD for assistance with statistical analysis.

